# Nurses’ Perceptions regarding Current Skills in Minimizing Patient’s Aggression at a selected Psychiatric hospital in Lesotho

**DOI:** 10.1101/2022.01.03.22268657

**Authors:** Libuseng Moureen Rathobei, Isabel Nyangu, Makhosazane Barbara Dube

## Abstract

**Background:** Meta-analysis of international literature indicate high frequencies of aggression in mental health setting. Several studies indicate that among professional health workers, nurses are more likely than other staff members to experience aggressive incidences from patients. Furthermore, based on cause, nurses apply range of interventions in managing aggression, however, despite their perceived value of their intervention in managing aggression, no single intervention is sufficient for handling aggressive patients to stay in control in wards. Despite high priority placed on patient aggression management, insufficient research studies have been conducted on nurses’ perceptions regarding current skills in minimizing patient’s aggression.

**Objectives:** The purpose of the study was to describe nurses’ perceptions regarding current assessment skills in minimizing patient’s aggression at selected Psychiatric hospital.

**Methodology:** Non-experimental, exploratory, descriptive research design was used to guide research process. All-inclusive sampling method was used with 119 nurses as respondents. Data was collected by means of a questionnaire, analyzed using SPSS version 25. Descriptive statistics procedure was used to present findings of the study.

**Results:** Minority of respondents, (n=27) 22.7% agreed that their current assessment skills for minimizing patients’ aggression were good compared to majority of respondents, (n=92) 77.3%, who expressed a different opinion. This suggests that there is limited current assessment skills in identifying high risk aggressive patients in a selected Psychiatric hospital.

**Conclusion:** The findings of study indicate that there is limited current assessment skills in minimizing patients’ aggression at selected Psychiatric hospital. Therefore, there is need to provide comprehensive information on nurses’ current skills in minimizing patients’ aggression to obtain richer information.

## Introduction/Background

Patients’ aggression in mental health care setting is global health problem with major psychological, physical and economic consequences (Ramezani, Gholamzadeh, Torabizadeh, Sharif and Ahmadzadeh, 2017). Several studies indicated that among professional health workers, nurses are more likely than other staff members to experience aggressive incidences from patients and visitors (Bekelepi, Martin and Chipps, 2015a; Baby, Gale and Swain, 2019). Shah, Annamalai, Aye, Xie, Ng, Shah and Manickam (2016) asserted that aggressive incidents refers to both physical and verbal abuse, threatening gestures, assaults and any other behavior directed at an individual that comprises his or her safety. Consequently, aggression prevention should be priority as it has huge impact on quality of care (Hallett and Dickens, 2017; Tölli, Partanen, Kontio and Häggman□Laitila, 2017).

Baby, Gale and Swain (2018a) stated that aggressive behaviors may disrupt therapeutic relationship if not addressed appropriately. Baby et al. (2019) stated that studies proffer staff-patient interactions as significant precursor to aggressive incidents especially in mental health-settings; negative staff interaction styles and limited communication skills are strong precursors of aggression. Therefore, based on the cause, nurses apply range of interventions in managing aggression, however, despite their perceived value of their interventions in managing aggression, no single intervention is sufficient for handling aggressive patients to stay in control in the wards (Ramezani et al., 2017). It is therefore necessary that nurses acquire necessary skills to manage aggressive patients (Baby, Gale and Swain, 2018b).

Price, Baker, Bee, Grundy, Scott, Butler, Cree and Lovell (2018), argue that serious service user concerns about use of physical restraint to manage aggression in psychiatric settings has serious health consequences which resulted in prioritization of non-physical approaches. According to Heckemann, Zeller, Hahn, Dassen, Schols and Halfens (2015) management of aggression include initiating and maintaining nurse-patient relationship. Furthermore, Guay, Goncalves and Boyer (2016) stated that it is recommended that employers implement Education and Training Programs in order to prevent aggression to help nurses develop skills to better recognize, react to aggressive situations and better cope with their consequences.

The study conducted in nursing college, Bangalore on 71 nursing students, results showed that level of knowledge and confidence was viewed as significantly higher during the post-test and comparison of knowledge level (t=13.2, p=0.00) and confidence level (t=5.58, p=0.00) which showed that aggression management training programs were effective on competency of nursing students. In contrast, study conducted in Psychiatric hospital, Nigeria of which objective was to facilitate participants’ descriptions of current experiences and practices towards aggression and its management, results indicated that participants felt betrayed by role players within Mental Health Care Service System; disappointed that de-escalation techniques were considered evidence-based practice and hopeless about introduction of de-escalation techniques as they perceived that it did not bear effective results in care of aggressive patients (Oyelade, Smith and Jarvis, 2017). Though, the study conducted by Bekelepi, Martin and Chipps (2015b) in Psychiatric hospital in Western Cape, South Africa, on 149 professional nurses, results indicated that that nurses with less years of experience felt skilful and confident than their counterparts who had more years of experience, however those who had training in aggression management perceived that training did not meet their needs and overall findings revealed that nurses had fair knowledge and skills in managing aggressive Psychiatric patients (Bekelepi et al., 2015b).

Management of aggression can be context bound, as what works in one situation may not work in another situation (Bimenyimana, Poggenpoel, Temane and Myburgh, 2016). Currently, there is no Evidence Based Research regarding nurses’ perceptions regarding current skills in minimizing patient’s aggression at selected Psychiatric hospital in Lesotho. However, anecdotal evidence and insight through work experience indicate that nurses have challenge in managing aggressive patients in Psychiatric hospitals in Lesotho without being hurt in process. Therefore, proposed study seeks to explore nurses’ current skills in managing patients’ aggression at a selected Psychiatric hospital in Lesotho.

### Conceptual Framework

The variables for the conceptual framework was based on Duxbury (2002) three-dimensional understanding of the causes of inpatient aggression, nurses’ skills in minimizing aggression and management strategies and Abderhalden, Needham, Friedli et al. (2002) descriptions of how nurses perceive inpatient aggression. Nurses generally perceive inpatient aggression in one of two ways: as functional and therefore understandable e.g. a form of communication, an anxiety management strategy; the start of the nurse-patient relationship; an attempt to protect oneself; or as dysfunctional and therefore unnecessary e.g. unacceptable; a negative response; an act of physical aggression against a nurse).

Duxbury (2002) three-dimensional model says aggression in inpatient settings occurs as a result of three broad interacting components namely the internal component, the external component and the situational/interactional component (Figure: I).

**Figure 1:**
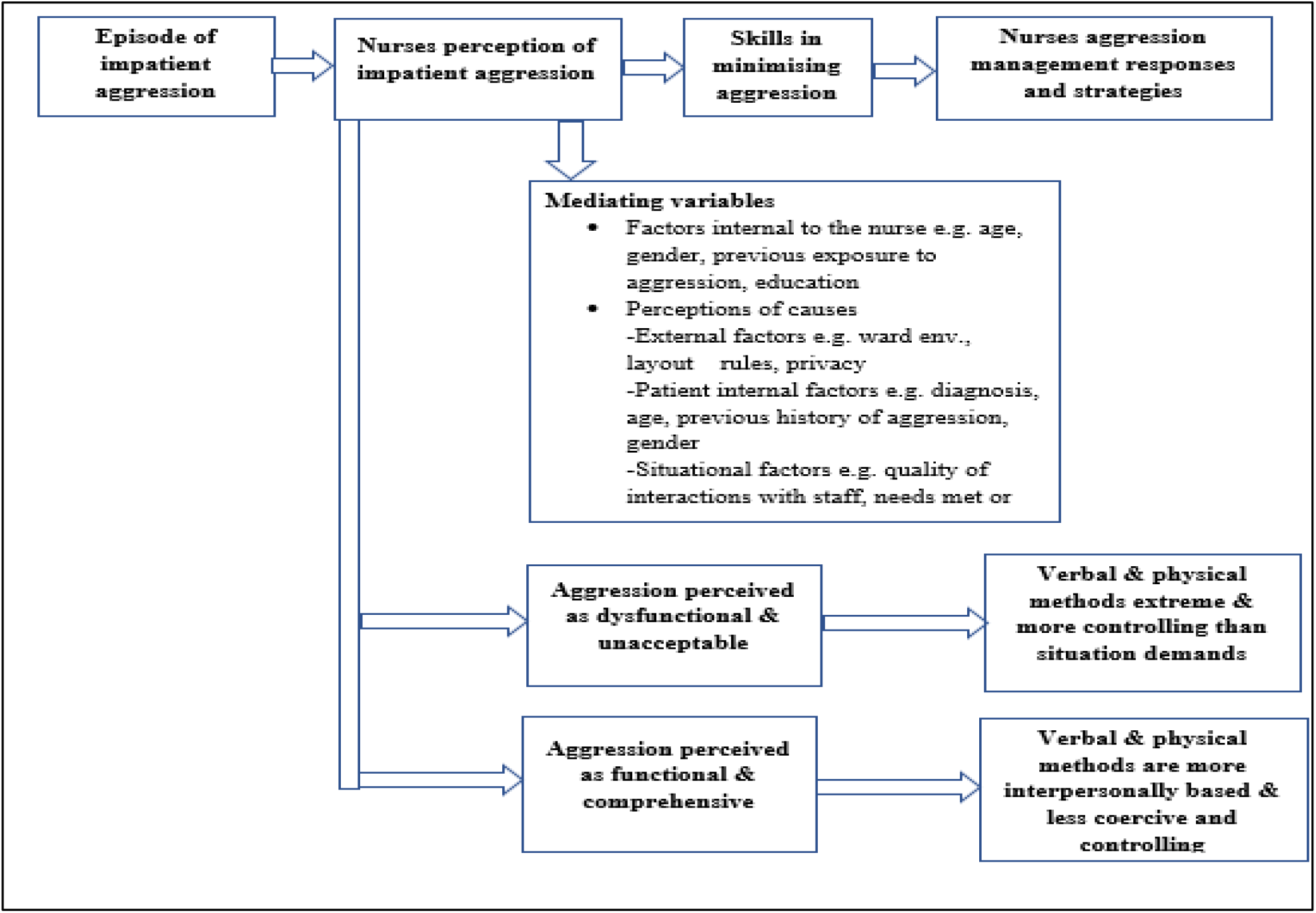
Conceptual framework (Adapted from Abderhalden, Needham, Friedli et al. (2002); Duxbury (2002)and (Ajzen, 1988) theory of planned behavior)

Internal model has been defined as those factors residing within the aggressive person, such as mental illness e.g. Schizophrenia, or personality Duxbury (2002). Age and gender have been associated with increased risk for aggression; men between the ages of 15-35 tend to be more at risk for aggression than younger or older men and women. The external model sees aggression being mainly caused by factors in the person’s physical or social environment, such as the physical layout of the ward, or the way in which the ward is governed by the staff. The situational/interactional model includes factors in the immediate situation, such as the interaction between the patient and others (particularly staff members). Nurses are likely to perceive these models differently, and their beliefs about causation may influence their approach to managing an aggressive incident (Foster et al., 2007).

This conceptual framework is underpinned by the theory of planned behavior which suggests that an individual’s perceptions of events (negative or positive) influence the nature of the skills he/she will take in response to those events (Ajzen, 1988). However, these perceptions and responses are mediated through a range of environmental factors (e.g. support from others in the environment for the intended action) personal characteristics (e.g. amount of experience with the phenomenon, problem-solving ability, degree of perceived personal control and mastery (Jonker et al., 2008; Nakahira, Moyle, Creedy et al., 2009; Van Wijk, 2006). The study therefore assumes that if the nurse perceives aggression as a dysfunctional and unnecessary behavior, he/she is more likely to use coercive strategies in managing this behavior Duxbury (2002). However, the nurses’ perceptions, skills and hence management will be mediated by her views on the cause of the aggression (internal to the client, located in the environment or within the interaction) and a range of personal characteristics such as level of education, nursing experience, age and gender and experience with violent behavior (Nakahira et al., 2009).

## Aims and Objectives

The purpose of the study was to describe nurses’ perceptions regarding current assessment skills in minimizing patient’s aggression at selected Psychiatric hospital.

## Methodology

A quantitative, non-experimental, exploratory and descriptive design was used to conduct research. Exploratory design studies full nature of phenomenon, the way it manifests and other factors that are linked with underlying process (Polit and Beck, 2016). According to Burns and Grove (2010), descriptive design maybe used for development of theory, identification of problems within current practice, modifying existing practice, making judgments or spotting what others in parallel situations are doing. This design was appropriate to attain information that describes “what exists” with reverence to nurses’ perceptions regarding current skills in minimizing patients’ aggression at a selected Psychiatric hospital in Lesotho.

### Setting

The study was conducted at selected Psychiatric hospital in Lesotho. The hospital provides specialized Psychiatric referral services for all district hospitals in country. It serves as clinical area for Mental Health placement for students from Psychology, General nursing; Diploma and Degree, Social work and Occupational Therapy students.

### Sampling

The population of the study was all nurses, 133, who worked and had exposure to aggressive patients in Psychiatric hospital from 2017. The population (n=133) included 15 psychiatric nurses; 16 registered nurses, 88 nursing students and 14 nursing assistants. Quota sampling was used to conveniently select respondents who completed a self-administered questionnaire. Data collection occurred over in September 2018. A total of 119 out of 133 questionnaires were returned completed and response rate was 89%.

### Data Analysis

Descriptive Analysis was used to describe and summarize nurses’ perceptions of types of aggressive behavior displayed by patients. According to Polit and Beck (2016), descriptive statistics are used sometimes to directly address research questions in studies that are primarily descriptive and help set stage for understanding of quantitative research evidence.

The questionnaire was developed by researcher with assistance of research supervisor and statistician using literature to guide development thereof. Five responses on a likert scale were; Strongly Disagree, Disagree, Uncertain, Agree and Strongly Agree. Face validity was established by consulting experts in the field of psychiatric nursing, the supervisor and statistician to provide feedback regarding validity of questionnaire. In order to maintain test-re-test reliability of the questionnaire, it was pre-tested in ten respondents and as no changes were made to the questionnaire, their data were included in the final sample. By means of Cronbach’s Alpha reliability statistics, validity of the instrument (questionnaire) was grounded 0.85 (85.0%) which entails that findings from this study would be reliable. The completed questionnaire were counted and coded to facilitate capturing and auditing of data after collection. Next, data was entered into computer Soft-ware Statistical Package for Social Science (SPSS) version 25 to analyze data. Descriptive statistics was used to describe and synthesize data where frequencies, percentages, standard deviations and means were reflected. Tables were used to enhance interpretation. Composite score was computed for questions with ranges in the questionnaire

### Ethical Consideration

Grove, Burns and Gray (2014) opine that researcher must comply with three ethical principles, namely; beneficence, respect for human dignity and justice. Permission to conduct the study was obtained from the Research Committee of the University of KwaZulu Natal (ID-HSS/067/018M), Lesotho Ministry of Health (ID-110/2018), and Nursing Service Manager of the selected psychiatric hospital. To respect rights of respondents, researcher explained purpose of study to all nurses and explained that participation in study was voluntary and they have right to withdraw at any time if they felt uncomfortable without fear of any negative effects. Researcher also explained that respondents would experience no harm by participating in study, however, negative perceptions of aggressive incidents and painful memories can be traumatic. Therefore, a contingency plan was put in place whereby the respondents who would experience emotional or psychological problems, would be referred to professionals at the hospital.

After providing necessary information regarding study, a signed informed consent was obtained from those who voluntarily accepted to participate. Researcher explained to respondents that questionnaire would take 20 minutes of their time to complete and that their anonymity and confidentiality was respected by using codes on questionnaires. Their names did not appear anywhere on questionnaire, so no one was able to identify the owner of response. Respondents were treated equally, and data was presented as it was collected, without modification. The data was kept safely in locked area which only researcher and supervisor had access. Researcher explained that findings of study would be helpful to nurses, including other nurses in general as they will have knowledge. The results of study may be useful in informing policies and review of policies in hospital, together with review of curriculum in schools of nursing.

## Results

A total of 133 questionnaires were handed out to total population of 133 nurses, of whom 119 completed and returned questionnaires. Nurses had varied perceptions on current skills in minimizing patients’ aggression at a selected Psychiatric hospital in Lesotho.

### Demographic information

The minimum age of respondents was 20 years, while the maximum age was 56 years old. The median was 27 years old, whereas the mode was 21 years old. The mean was 29.8 years (±9.2). Pearson Chi-square test was performed to see the association between age and nurses’ current skills in minimizing patient’s aggression and there was a statistically significant association (X^2^=20.637, d.f = 3 and p.000)

**Figure 3:**
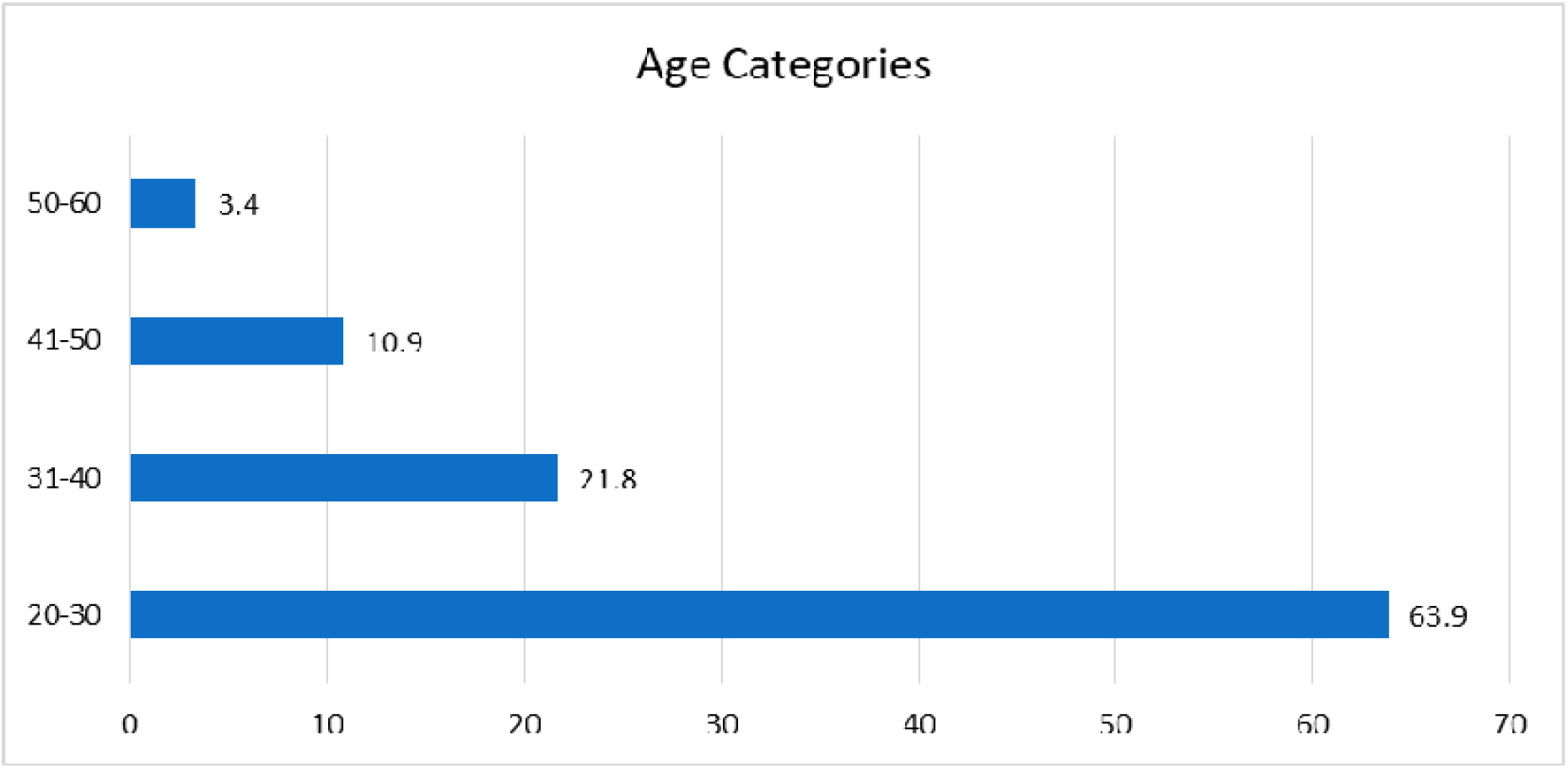
Bar Chart Showing Respondents Age.

Majority of respondents, 71.7% (n=85) were females, while 28.6% (n=34) were males. Pearson Chi-Square test was performed to observe the association between gender and current skills in minimizing patients’ aggression and there was statistically significant association (X^2^=25.967, d.f= 3 and p=0=.000).

**Figure 4:**
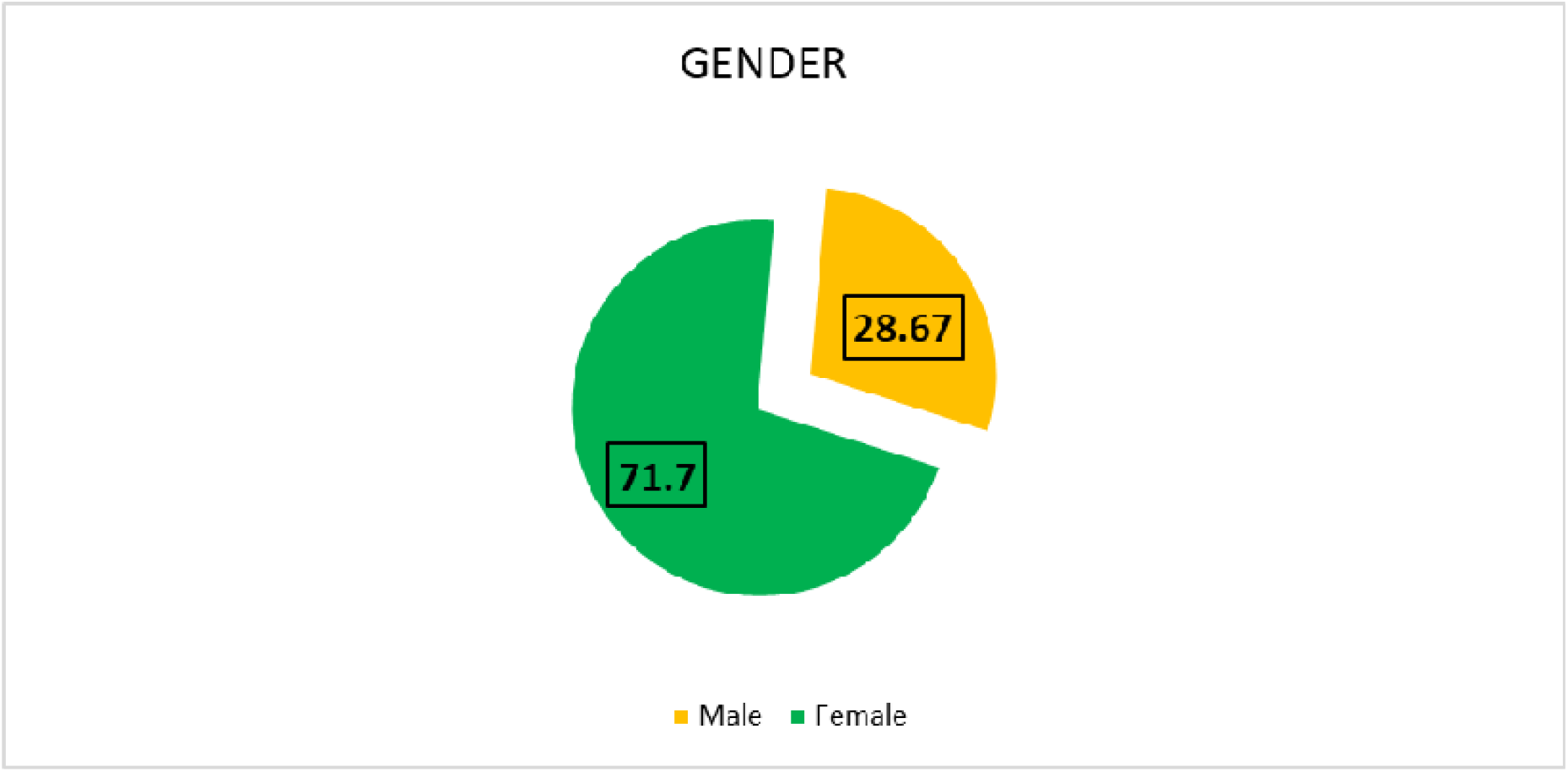
Gender of the respondents (n=119)

Of all nurses, 67.2% (n=80) were student nurses, 10.1% (n=12) trained nursing assistants, 11.8% (n=14) registered nurses and 10.9% (n=13) registered psychiatric nurses. The number of years of experience was less than one year for 67.2% (n=80) of respondents, six to 10 years for 13.4% (n=16), one to five years for 9.2% (n=11), 11-20 years for 7.6% (n=9) and 20-30 years for 2.5% (n=3). Pearson Chi-square test was performed to observe the association between years of experience and nurses’ current skills in minimizing patients’ aggression and there was statistically significant association (X2=20.209, d.f=4 and p=.000).

**Figure 5:**
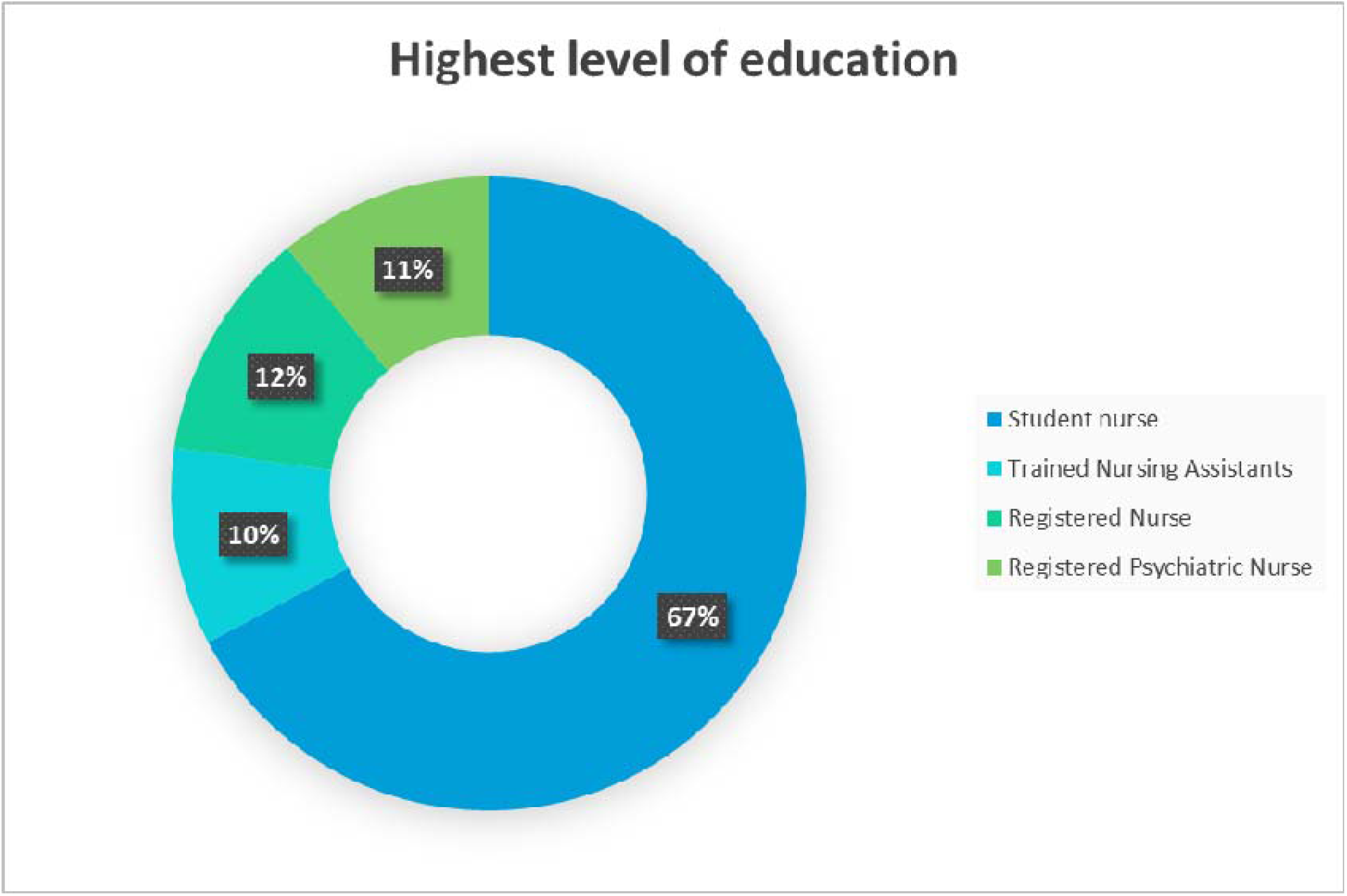
Highest Level of Education.

### Current skills in minimizing patients’ aggression

When exploring the statement on, “*My current assessment skills for identifying high risk aggressive patients are good”*, minority of respondents, 3.4% (n=4) strongly agreed and 8.4% (n=10) agreed to the statement, while most of the respondents, 53.8% (n=64) strongly disagreed and 25.2% (n=30) disagreed, whereas 9.2% (n=11) of the respondents felt uncertain on the statement that their current assessment skills for identifying high risk aggressive patients are good.

On the notion that, “*My current skills in identifying early aggression warning signs are good’*, minority of the respondents, 4.2% (n=5) strongly agreed and 8.4% (n=10) agreed to the statement, while majority of the respondents, 44.5% (n=53) strongly disagreed and 31.1% (n=37) of the respondents disagreed. However, 11.8% (n=14) of the respondents reported not sure if their current skills in identifying early aggression warning signs are good.

On the statement that, “*I am aware of the cycle of aggression and responses to use at each stage”*, minority of the respondents, 4.2% (n=5) strongly agreed and15.1% (n=18) agreed to the statement that they are aware of the cycle of aggression and responses to use at each stage, while 31.9% (n=38) of the respondents strongly disagreed and 32.8% (n=39) disagreed, whereas, 16.0% (n=19) of the respondents felt uncertain about whether they are aware of the cycle of aggression and responses to use at each stage.

Based on the statement that, “*I am aware of different options to use when confronted with aggression”*, out of 100.0% (n=19) of the respondents, minority of them, 8.4% (n=10), strongly agreed and 26.9% (n=32) agreed, while majority of respondents, 29.4% (n=35) strongly disagreed and 21.0% (n=25) disagreed to the statement that they are aware of different options to use when confronted with aggression. 14.3% (n=17) of the respondents felt not sure on whether they are aware of different options to use when confronted with aggression.

Regarding the notion that, “*I am able to use verbal and non-verbal skills for handling aggression*”, minority of respondents, 16.0% (n=19) strongly disagreed, 16.0% (n=19) disagreed to the statement, while majority of the respondents, 32.8% (n=39) agreed and 19.3% (n=23) strongly agreed to the statement that they are able to use verbal and non-verbal skills for handling aggression. However, 16.0% (n=19) of the respondents felt uncertain on whether they are able to use verbal and non-verbal skills for handling aggression.

Based on the statement, “*I am able to use physical break away skills if I am physically attacked*”, there were no respondents who strongly agreed to the statement and minority of the respondents, 3.4% (n=4) of the respondents agreed to the statement, while majority of the respondents, 58.0% (n=69) strongly disagreed and 27.7% (n=33) of the respondents disagreed, whereas, 10.9% (n=13) of the respondents reported not sure if they are able to use physical break away skills if they are physically attacked, with the lowest mean score of 1.60.

On the fact that, “*I am able to use team skills to restrain a patient who is aggressive*”, majority of the respondents, 29.4% (n=35) strongly agreed and 29.4% (n=35) of the respondents agreed, whereas minority of the respondents, 19.3% (n=23) strongly disagreed and 7.6% (n=9) disagreed to the statement that they are able to use team skills to restrain a patient who is aggressive. 14.3% (n=17) of the respondents felt uncertain on whether they are able to use team skills to restrain a patient who is aggressive, with the highest mean score of 3.42 and coefficient of standard deviation of 0.49.

Based on the statement that, “*I am able to use de-escalation when encountering an aggressive patient”*, minority of the respondents, 0.8% (n=1) strongly agreed and 2.5% (n=3) agreed while majority of the respondents, 57.1% (n=68) of the respondents strongly disagreed and 29.4% (n=35) of the respondents disagreed to the statement that they are able to use de-escalation when encountering an aggressive patient, whereas 10.1% (n=12) of the respondents reported not sure if they were able to use de-escalation when encountering an aggressive patient.

Regarding the statement that, “*I am aware of my legal rights and limits when defending myself against aggression”*, majority of the respondents, 23.5% (n=28) strongly agreed and 19.3% (n=23) agreed to the statement, while minority of the respondents, 30.3% (n=36) strongly disagreed and 11.8% (n=14) of the respondents disagreed to the statement that they are aware of their legal rights and limits when defending themselves against aggression. 15.1% (n=18) of the respondents felt not sure if they are aware of their legal rights and limits when defending themselves against aggression.

Based on the fact that, “*I am aware of the procedures for reporting and documenting aggressive incidents in my work*”, majority of respondent, 26.1% (n=31) strongly agreed and 18.5% (n=22) of the respondents agreed, while minority of the respondents, 27.7% (n=33) strongly disagreed and 12.6% (n=15) of the respondents disagreed to the statement that they are aware of the procedures for reporting and documenting aggressive incidents in their work, whereas 15.1% (n=18) of the respondents were uncertain on whether they are aware of the procedures for reporting and documenting aggressive incidents in their work.

Based on the notion that, “*I am aware of the options and procedures for debriefing after aggressive incidents in my workplace”*, minority of the respondents, 11.8% (n=14) strongly agreed and 22.7% (n=27) of the respondents agreed, while majority of the respondents, 35.3% (n=42) strongly disagreed and 10.9% (n=13) of the respondents disagreed to the statement that they are aware of the options and procedures for debriefing after aggressive incidents in their work. 19.3% (n=23) of the respondents felt not sure whether they are aware of the options and procedures for debriefing after aggressive incidents in their workplace.

## Discussion of Results

When exploring whether the respondents’ current assessment skills for identifying high aggressive patients are good, the study indicated that majority, 53.8% of the respondents strongly disagreed with the statement that their current assessment skills for identifying high aggressive patients were good. This may be due to lack of evaluation of their different perceived assessment skills for identifying high aggressive patients. This is in contrast with the study conducted by Bekelepi,et al (2015) who stated that majority of the respondents reported that they were able to assess the aggressive patients. This is also in contrast with the study conducted by Partridge and Affleck (2018), where it was discovered that majority of nurses used Broset Violence Checklist to identify high risk aggressive patients.

The study showed that majority of the respondents, 44.5% strongly disagreed with the statement that their current skills for identifying early aggression warning signs are good. This may be due to nurses’ minimum confidence in their current skills for identifying early aggression warning signs. This is in contrast with the study conducted by Partidge and Affleck (2018) where it was discovered that majority of nurses used Broset Violence Checklist to identify high risk aggressive patients early. In addition, Fluttert, et al (2011) asserted that early signs of verbal aggression can be defined as the subjective perceptions, thoughts and behaviours of the patient occurring prior to the incident of violence or aggressive behaviour and therefore need to be identified. Furthermore, Uys and Middleton (2014) opined that it is not possible to predict whether a particular patient will become violent in the future, but it is possible to predict aggression in short-term.

The results of the study indicated that majority of the respondents, 32.8% disagreed with the statement that they were aware of the cycle of aggression and responses to use at each stage. This may be due to escalation of aggression in the psychiatric hospital. This study is in line with Casey (2019), who stated that aggression in psychiatric hospitals towards nurses has increased. The study is in keeping with the study conducted by Chang, Park, Jang, Ahn and Yoon (2019), who stated that 70% of nurses had experienced at least 1 type of verbal aggression whereby higher physical aggression and hostility were significantly related to greater verbal abuse experience. Furthermore, Halm (2017), echoed this sentiment, in the study conducted in Australia which stated that the percentage of nurses reporting having experienced verbal threats or actual aggression ranges from 33% to 65%.

Based on the fact that the respondents were aware of different options to use when confronted with aggression, most of the respondents, 29.4% strongly disagreed with the statement that they were aware of different options to use when confronted with aggression. This may be due to heightened perception on the use of physical restraints. This study is in line with the study conducted by Eskandari, Abdullah, Zainal and Wong (2017) on 309 nurses, whereby the results revealed that less than half of nurses considered alternatives to physical restraint and that multiple linear regression analysis found knowledge, attitude and intentions were significantly associated with nurses’ practice to use physical restraint. This study is in contrast with McCann et al. (2014), who stated that clinical staff use a range of personal-centred containment (e.g. restraint, seclusion, medication) measures to manage their behaviours when confronted by aggression.

The results of the study showed that most of the respondents, 32.8% agreed that they were able to use verbal and non-verbal skills for handling aggression. This may be due to nurses’ communication skills which are taught during their training. This is in line with the study conducted by Bekelepi et al. (2015), who stated that majority of the respondents reported that they use verbal and non-verbal communication with patients. This is also congruent with Riley (2015), who posit that there are two parts of face to face communication; verbal expression of the sender’s thoughts and feelings and non-verbal expressions. However, the current study is in contrast with Shafakhah et al. (2015), who asserted that despite the effect of communication skills on the quality of nursing care and patient improvement and participation in care, the results showed that nurses were not successful in communicating (using verbal and non-verbal) with patients and families, nurses communicated with patients for a very short period of time. This is also in contrast with Papageorgiou et al. (2017), who stated that nurses find it difficult to communicate effectively with patients with severe mental illness about symptoms, drug treatment, and their side effects and to reach a shared understanding about the diagnosis, prognosis and treatment. The study is also contrary to the study conducted by Shafakhah et al. (2015), who demonstrated that most nursing students required improvements in their communication skills (both verbal and non-verbal) in both clinical communication behaviour and treatment communication ability.

Majority of the respondents, 58.0% strongly disagreed with the statement that they were able to use physical breakaway skills, if they are physically attacked. This may be as a result of lack of institutional training on physical breakaway skills and on lack of knowledge on physical breakaway skills. The current study is congruent to Harwood (2017) who asserted that in physical breakaway techniques, nurses need to ensure that their escape route is clear in case he or she has to withdraw quickly from aggressive patients.

The study suggested that 29.4% of the respondents strongly agreed and 29.4% of the respondents agreed that they were able to use team skills to restrain a patient who is aggressive. This may be due to the skills taught during their training on managing mentally ill patients. This is in engagement to the study conducted by Bekelepi, et al (2015) who asserted that majority of the respondents reported that they were able to use team skills to restrain patients. This is also in line with Palaganas, et al, (2014) who stated that implementation of structured team approach with good communication that promotes inter-professional collaboration to manage patients with aggression has shown significant impact on mitigating aggression by patients.

The study results showed that majority of respondents, 57.1% strongly disagreed that they were able to use de-escalating techniques when encountering an aggressive patient. This may be due to lack of training on de-escalation techniques. The results of this study are in contrast with the study conducted by Baig, Tanzil, Shaikh, Hashmi, Khan and Polkowski (2018) among healthcare providers in Karachi, whereby 147 healthcare workers reported to have received trainings on de-escalation and reported to be effective with the overall assumption of 20% reduction in the frequency of aggression. This study is also contrary to the study conducted by Price et al. (2018), to obtain staff descriptions of de-escalation techniques currently used in mental health setting, nurses reported using de-escalation techniques in response to escalated aggression.

In regard to the statement that the respondents were aware of their legal rights and limits when defending themselves against aggression, majority of the respondents, 30.3% strongly disagreed. This may be due to their knowledge regarding health care law and rights of mentally ill patients. This study is in contrast to the study conducted by Bekelepi et al. (2015), who stated that majority of the respondents reported that they were able to use policies that regulate physical restraints and seclusion. Furthermore, McCann et al. (2014), stated that according to legal rights and limits, seclusion and restraint are contrary to prominent International recommendations, Government reports, mental Health Service Policies, which advocates that as strategies to deal with disturbed behaviour, restraint and seclusion must be used as little as possible or eliminated. In addition, Hallett and Dickens (2015) opined that the National Clinical Guidelines state that coercive interventions including seclusion, restraint and rapid tranquilization should only be considered once de-escalation strategies have failed.

On the statement that the respondents were aware of the procedures for reporting and documenting aggressive incidents at work, majority of the respondents, 27.7% strongly agreed. This is in line with Campbell et al. (2015), who indicated that despite the current National and International attention focused on patient aggression in healthcare setting, patient aggression towards healthcare workers remains under-reported. Furthermore, this is congruent to Arnetz et al. (2015), who posted that under-reporting of aggressive events has been identified as a failure of victimized employees to report to their employers or their officials and is the under-estimation of the true extent of the problem, thus indicating less of the need for prevention of possible negative effects that may actually be warranted. However, the study is in contrast to the study conducted by Bekelepi et al. (2015), who stated that majority of the respondents reported that they are able to follow procedure for reporting and documenting incident of aggression.

In regard to the statement that the respondents were aware of the options and procedures for debriefing after aggressive incidents in their work, 35.3% of the respondents strongly disagreed. This is in line with Kessler et al. (2015), who posited that in a meta-analysis of a team-based debriefing after clinical events, there was improved effectiveness in teams that debriefed compared with those that did not.

**Table 14:** Nurses’ Current skills in Minimizing Patient’s Aggression.

| Variable | SD |  | D |  | U |  | A |  | SA |  | Mean |
| --- | --- | --- | --- | --- | --- | --- | --- | --- | --- | --- | --- |
|  | Freq | % | Freq | % | Freq | % | Freq | % | Freq | % |  |
| My current assessment skills for identifying high risk aggressive patients are good | 64 | 53.8 | 30 | 25.2 | 11 | 9.2 | 10 | 8.4 | 4 | 3.4 | 1.82 |
| My current skills in identifying early aggression warning signs are good | 53 | 44.5 | 37 | 31.1 | 14 | 11.8 | 10 | 8.4 | 5 | 4.2 | 1.97 |
| I am aware of the cycle of aggression and responses to use at each stage | 38 | 31.9 | 39 | 32.8 | 19 | 16.0 | 18 | 15.1 | 5 | 4.2 | 2.27 |
| I am aware of different options to use when confronted with aggression | 35 | 29.4 | 25 | 21.0 | 17 | 14.3 | 32 | 26.9 | 10 | 8.4 | 2.64 |
| I am able to use verbal and non-verbal skills for handling aggression | 19 | 16.0 | 19 | 16.0 | 19 | 16.0 | 39 | 32.8 | 23 | 19.3 | 3.24 |
| I am able to use physical break away skills if I am physically attacked | 69 | 58.0 | 33 | 27.7 | 13 | 10.9 | 4 | 3.4 | 0 | 0.0 | 1.60 |
| I am able to use team skills to restrain a patient who is aggressive | 23 | 19.3 | 9 | 7.6 | 17 | 14.3 | 35 | 29.4 | 35 | 29.4 | 3.42 |
|  | Freq | % | Freq | % | Freq | % | Freq | % | Freq | % |  |
| I am able to use de-escalation when encountering an aggressive patient | 68 | 57.1 | 35 | 29.4 | 12 | 10.1 | 3 | 2.5 | 1 | 8 | 1.61 |
| I am aware of my legal rights and limits when defending myself against aggression | 36 | 30.3 | 14 | 11.8 | 18 | 15.1 | 23 | 19.3 | 28 | 23.5 | 2.94 |
| I am aware of the procedures for reporting and documenting aggressive incidents in my work | 33 | 2.7 | 15 | 12.6 | 18 | 15.1 | 22 | 18.5 | 31 | 26.1 | 3.03 |
| I am aware of the options and procedures for debriefing after aggressive incidents in my workplace | 42 | 35.3 | 13 | 10.9 | 23 | 19.3 | 27 | 22. | 14 | 11.8 | 2.65 |

### Limitation of the study

- The researcher only collected data from nurses using self-reported questionnaires
- The study was only localized at referral Psychiatric hospital, therefore the findings cannot be generalized to other Psychiatric institutions

### Recommendations

This study focused on Quantitative approach that used questionnaire for data collection. It is recommended that qualitative study should be conducted on the same topic to obtain richer data on the nurses’ perceptions regarding current skills in minimizing patients’ aggression at a selected Psychiatric hospital.

## Conclusion

The findings of research indicate that nurses perceive their current assessment skills in minimizing patients’ aggression limited. Therefore, there is need to provide current skills to nurses that will minimize patient’s aggression at a selected psychiatric hospital.

## Data Availability

All data produced in the present work are contained in the manuscript

